# Student attributes and behavioral examples used by physiotherapy clinical educators in Hong Kong: A qualitative data analysis

**DOI:** 10.64898/2026.01.13.26344005

**Authors:** Doris Y.K Chong, Catherine M. Capio, Alice Y.M. Jones, Percy P.S. Tse, Sammi H.Y. Chau, Kathlynne F. Eguia

**Author notes:** Corresponding author: Doris Y.K Chong, Associate Professor Department of Physiotherapy, School of Nursing and Health Sciences Hong Kong Metropolitan University, Hong Kong SAR. **Author contributions: CRediT**. Doris Y.K. Chong: conceptualization, data curation, formal analysis, funding acquisition, investigation, methodology, project administration, supervision, validation, writing-original draft, writing - review and editing. Catherine M. Capio: conceptualization, funding acquisition, investigation, methodology, validation, writing - review and editing. Sammi H.Y. Chau: data curation, funding acquisition, writing - review and editing Alice Y.M. Jones: validation, writing - review and editing. Percy P.S. Tse: data curation, formal analysis, methodology, software, writing-original draft, writing - review and editing. Kathlynne F. Eguia: data curation, formal analysis, methodology, software, validation, writing-original draft, writing - review and editing.

## Abstract

**Background:** Clinical placements are a vital element of physiotherapy education, where students must demonstrate competence across a range of professional attributes. Although core competencies such as clinical knowledge, ethical conduct, and communication are universally valued, clinical educators (CEs) from different cultural contexts may emphasize these attributes in different ways. Gaining insight into how Hong Kong CEs evaluate students is important for aligning academic expectations with clinical practice.

**Objective:** This study identifies the key student attributes prioritized by Hong Kong CEs and summarizes the behavioral examples they use to distinguish performance levels on the Assessment of Physiotherapy Practice (APP) Global Rating Scale.

**Methods:** A secondary qualitative analysis was conducted on 456 qualitative feedback comments from APP forms completed by 45 CEs assessing physiotherapy students across two cohorts. The data were analyzed using AI-assisted thematic analysis combined with human expert interpretation, followed by deductive validation across performance levels (Excellent, Good, Adequate, Not Adequate).

**Results:** Six core attributes emerged from the analysis: (1) communication and interpersonal skills, (2) clinical reasoning and decision-making, (3) practical knowledge and technical competence, (4) learning attitudes and reflective practice, (5) professionalism and work ethics, and (6) safety and risk management, and patient-centered care. Behavioral examples were mapped across performance levels, revealing clear distinctions between competent and underperforming behaviors. Among these attributes, *learning attitudes and reflective practice* were consistently emphasized, reflecting cultural values within the Hong Kong clinical education context.

**Conclusion:** Hong Kong CEs prioritize not only technical and cognitive competence, but also reflective and affective attributes rooted in professional and cultural values. The identified attributes and behavioral descriptors may provide actionable guidance for curriculum design, educator training, and student preparation, fostering coherent, transparent, and culturally informed clinical assessment practices.

## Introduction

Clinical placements represent a critical component of physiotherapy education, typically comprising approximately one-third of the curriculum (World Physiotherapy, 2021). These immersive, hands-on experiences provide students with essential opportunities to apply their theoretical knowledge in real-world clinical settings (Thomson et al, 2014), thereby facilitating the development of essential practical competencies. During clinical placements, students are continuously assessed and receive feedback not only on their core physiotherapy knowledge and clinical skills but also on other key professional attributes, such as communication (Almeida Santos, Queirós, Couto, and Meneses, 2022; Chapman, Woo, and Maluf, 2022; Good, Lance, and Rainey, 2015). While many of the generic attributes expected of physiotherapy students, such as clinical competence, ethical and professional behavior and communication, are similar in many countries (Magni et al, 2025), clinical educators (CEs) with various cultural backgrounds may prioritize these attributes differently based on local cultural norms and expectations. This variation can create challenges in ensuring that both students and academic staff share a clear and consistent understanding of which attributes CEs pay close attention to in a specific cultural setting. Research has shown that although academics and CEs often share broad assessment criteria, they tend to prioritize and interpret these criteria differently when evaluating student performance (Cross, 1998). Therefore, identifying and clarifying the key qualities or behaviors that CEs notice and consider important during clinical placement is necessary to improve alignment between academic and clinical perspectives, allowing for more effective assessment practices and student learning experiences.

The Assessment of Physiotherapy Practice (APP), first established in Australia and now adopted internationally, is a reliable and validated tool used to assess physiotherapy students’ competence during clinical placements (Dalton, Davidson, and Keating, 2011). The APP consists of structured evaluation of the following 7 practice domains: professional behavior, communication, assessment, analysis and planning, intervention, evidence-based practice, and risk management. These domains are assessed through 20 items, each scored on a five-point scale ranging from 0 to 4. Additionally, students receive an ‘overall’ placement performance rating on the APP’s Global Rating Scale (GRS), which categorizes performance as *not adequate, adequate, good or excellent*. The APP has been adopted by one physiotherapy program in Hong Kong (HK) and is well regarded by local CEs (Chong, Chau, Capio, and Jones, 2024).

An essential element of the APP assessment process is the provision of qualitative feedback, which plays a critical role in learning. Feedback helps students identify discrepancies between their current and desired performance, providing motivation for improvement and reinforcing good clinical practice (Burgess and Mellis, 2015). CEs are encouraged to provide qualitative feedback that elaborates on students’ domain-specific scores and assists in interpreting the GRS score. This qualitative feedback not only enriches the quantitative APP scores but also facilitates better understanding of performance expectations by both students and educators.

A clear understanding of the expectations of CEs by both students and university educators is essential for the delivery of a successful clinical education program (Burgess, Van Diggele, Roberts, and Mellis, 2020). However, specific student clinician attributes prioritized by HKCEs and the behavioral examples they use to illustrate levels of student performance remain unclear in the Hong Kong context. This study aims to address this gap by exploring (i) which student attributes are most frequently observed and prioritized by HKCEs, as reflected in their qualitative feedback during placements, and (ii) what behavioral examples do HKCEs use in their feedback to characterize each performance level and whether these examples capture competent and underperforming behaviors associated with the GRS performance levels described in the APP. Previous studies have described feedback as specific information about observed performance in clinical education settings (van den Berg, Admiraal, and Pilot, 2006; Burgess and Mellis, 2015). For the purposes of this study, the phrase “most observed” refers to the attributes most reflected and emphasized in the qualitative, written feedback provided by HKCEs during clinical placements.

The findings from this study will contribute to the advancement of physiotherapy education in Hong Kong by facilitating a clearer understanding of the performance standards expected of students and informing the development of the physiotherapy curriculum to enhance preparation for pre-clinical placements. To achieve these aims, we conducted a secondary qualitative analysis of data from the APP generated during the clinical placements of physiotherapy students from Hong Kong Metropolitan University. The research questions and objectives are presented in Table 1.

**Table 1.** Research questions and objectives.

|  |  |
| --- | --- |
| **Research Question:** | "What student attributes do Hong Kong clinical educators (HKCE) prioritize during physiotherapy clinical placements, what behavioral examples from their feedback characterize each GRS performance level in the APP, and to what extent do the behavioral examples they described differentiate between GRS performance levels in the APP?" |
| **Research Objective 1:** | To identify and describe the key attributes and behavioral examples of student performance in clinical placements from the perspectives of CEs based on the qualitative feedback component of the APP. |
| **Research Objective 2:** | To examine what behavioral examples from HKCE feedback characterize each GRS performance level and to what extent the behavioral examples differentiate between GRS performance levels during clinical placements. |

## Methods

### Study Design, Participants and Data Collection

This study adopted a qualitative research design using secondary analysis of existing CEs’ qualitative feedback from APP forms collected during physiotherapy students’ clinical placements to identify key attributes of competence. The qualitative approach was deemed appropriate, as it enables in-depth exploration of complex educator perceptions and language that are not easily quantifiable (Cleland, 2017). Secondary data analysis enables the exploration of new research questions using existing rich textual data collected originally for educational assessment purposes and offers practical advantages in terms of resource efficiency (Cheong, Lyons, Houghton, and Majumdar, 2023; Johnston, 2014; Tripathy, 2013). The data were analyzed using thematic analysis, a qualitative method for identifying perspectives and themes related to feedback across various clinical education settings (Bearman et al, 2025; Mohd Noor et al, 2025).

Data were gathered from two cohorts (40 and 49 students) in an undergraduate physiotherapy program in HK (September 2023–May 2024 and September 2024–May 2025). During each period, the students completed five five-week clinical placements in neurological, musculoskeletal, cardiopulmonary, multisystem and community healthcare, separated by one-week breaks. Forty-five CEs from public hospitals, private hospitals, non-governmental organizations and private clinics, all formally trained in using the APP, supervised and assessed students (Chong, Chau, Capio, and Jones, 2024). Feedback written in the “final feedback box” of the APP form was retrieved for analysis and repurposed from routine educational assessment for this secondary qualitative study.

### Data Preparation

All personally identifiable information was anonymized to protect confidentiality and ensure data integrity. A total of 456 end-unit APP forms containing qualitative comments were collected for analysis. The comments were categorized by GRS scores as follows: Excellent (N=59), Good (N=271), Adequate (N=109), and Not Adequate (N=17). In this study, two AI-assisted tools (myRA Pro and ChatGPT-4.5) were incorporated into the qualitative analysis workflow to support coding and theme development. Because the AI-assisted analysis tool myRA can analyze only 20 transcripts per cycle, all 456 qualitative comments were randomly assigned (using a computer-generated random sequence) to 20 transcripts, each containing a mixture of GRS score categories to ensure representation across performance levels, and data files were prepared in formats compatible with both AI tools.

### AI-assisted Data Analysis

AI tools were used to enhance the efficiency and consistency of qualitative analysis when working with this large dataset. In line with the emerging applications of AI in general healthcare and physiotherapy research, these tools support the management of complex qualitative data, while human researchers retain responsibility for interpretation and judgment (Lan and Zhou, 2025; Perkins and Roe, 2024b; Reoli, Marchese, Duggal, and Kaplan, 2025; Zhang et al, 2025). Recent work further conceptualizes AI as a co-researcher that collaborates with humans across the qualitative research workflow, supporting both analysis and reflexive engagement with the data (Costa, Bryda, Christou, and Kasperiuniene, 2025). AI-assisted methods rapidly detect patterns and organize data, whereas researchers ensure methodological rigor and ethical integrity, which is consistent with prior work demonstrating that AI can accelerate thematic analysis but still requires expert oversight (Nicmanis and Spurrier, 2025; Perkins and Roe, 2024a; Williams, 2024). Using AI in thematic analysis may also facilitate identification of a broader range of themes, including subtle but meaningful patterns that might be missed by human analysis alone (Perkins and Roe, 2024a). Consistent with the literature, this study applies AI and human expertise in a complementary manner, with AI identifying potential themes and researchers providing critical reflection and judgment to refine the analysis (Perkins and Roe, 2024a, 2024b; Zhang et al, 2025).

### Data analysis

An inductive analysis was conducted using AI-assisted tools to identify emergent student attributes from CEs’ qualitative feedback without relying on preconceived categories. Following AI-assisted coding, investigators critically reviewed and validated the themes, engaging reflexively with the dataset to interpret prioritized attributes and identify any additional themes not captured by AI. The research team discussed and further refined the attributes to fit the context of the assessment.

A deductive analysis phase then tested whether these emergent attributes appeared consistently across different GRS performance levels (Excellent, Good, Adequate, Not Adequate), with separate data files prepared for each level. This analysis enabled validation of the robustness and generalizability of the attributes and the identification of behavioral examples that differentiated between GRS levels by matching educators’ feedback to the refined attribute categories. It helped assess the extent to which behavioral examples differentiate between GRS levels. Representative behavioral examples for each GRS level were then identified from the qualitative feedback, and the final attributes and examples associated with each rating were reviewed by the team to ensure relevance and consistency, with discrepancies resolved through discussion.

## Results

### Key Attributes

Analysis of 456 end-of-unit APP forms containing qualitative feedback from two cohorts of undergraduate physiotherapy students in Hong Kong identified six distinct attributes reflecting core aspects of student competence within the clinical environment. These attributes include (1) communication and interpersonal skills, (2) clinical reasoning and decision making, (3) practical knowledge and technical competence, (4) learning attitudes and reflective practice, (5) professionalism and work ethics, and (6) safety and risk management and patient-centered care. Table 2 outlines these key attributes alongside their corresponding descriptions, each carefully interpreted and finalized by the research team based on a synthesis of insights provided by both AI and human analysis of the feedback.

**Table 2.** Identified key attributes and their description.

| Key Attributes | Description | Sample quotes |
| --- | --- | --- |
| *Communication and Interpersonal skills* | Communication and interpersonal skills refer to the ability to clearly, empathetically, and effectively exchange information with patients, families, clinical educators, and healthcare team members. This includes using verbal and non-verbal communication appropriately, building rapport, actively listening, providing clear instructions and patient education, collaborating within multidisciplinary teams, and maintaining accurate and organized documentation. | *"She exhibited good communication skills with effective and efficient verbal guidance interspersed with demonstration..."* |
| *Clinical Reasoning and Decision Making* | Clinical reasoning and decision making refer to the thoughtful application of theoretical, evidence-based, and practical knowledge through critical thinking. Students systematically assess and interpret clinical information, identify and prioritize problems, develop appropriate treatment plans, adapt interventions based on patient responses, and justify clinical decisions logically. It reflects the cognitive processes behind effective patient management. | *"Student was able to conduct precise assessments, interpret clinical findings, and prioritize patient's problems appropriately."* |
| *Practical Knowledge and Technical Competence* | Practical knowledge and technical competence refer to the ability to effectively apply foundational theoretical knowledge in clinical practice through skilled hands-on performance. This includes conducting thorough patient interviews and objective assessments, safely delivering interventions and manual techniques, accurately performing procedures, and managing clinical workflows efficiently. It emphasizes practical proficiency, safe patient handling, and the integration of evidence-based treatments. | *"She is also required to get more familiar with the operation of ET modalities."* |
| *Learning attitude and reflective practice* | Learning attitude and reflective practice describe a proactive commitment to continuous personal and professional growth. This involves actively seeking feedback, showing openness to constructive criticism, engaging in honest self-assessment to identify strengths and areas for improvement, reflecting on clinical experiences, and adapting behavior accordingly. Students demonstrate eagerness, initiative, and motivation to pursue lifelong learning and to keep their clinical knowledge and skills current. | *"She is a bit passive in learning, cannot show her commitment or initiative to learn."* |
| *Professionalism and Work Ethic* | Professionalism and work ethic in physiotherapy involve consistent respectful, ethical, and mature behavior toward patients, clinical educators, and peers. These qualities are shown through punctuality, reliability, effective time management, and consistent attendance, reflecting strong commitment to clinical and academic responsibilities. Students demonstrate confidence, care, humility, and patience in their interactions, particularly with patients, while maintaining positivity, adaptability, and emotional stability to support effective teamwork and quality patient outcomes. | *"Student demonstrated very mature and professional behaviours throughout the placement."* |
| *Safety, Risk Management, and Patient-Centered Care* | Safety and risk management, coupled with patient-centered care, involve maintaining patient and practitioner safety through adherence to clinical standards and protocols while providing holistic, individualized care that respects each patient's unique preferences, needs, and values. Students are expected to identify, anticipate, and mitigate potential risks during patient care by ensuring safe assessment and treatment techniques. Students actively engage patients in their treatment and self-management, tailor interventions to individual abilities and circumstances, and prioritize patient comfort and safety in all clinical decisions. | *"He is advised to pay attention to safety awareness in positioning of patient and prescribing exercise."* |

### Behavioral Examples

Analysis of CEs’ narrative comments yielded a structured set of behavioral examples across six main domains mentioned above. Within each domain, comments were mapped to four global rating scale (GRS) levels (Excellent, Good, Adequate, and Not Adequate), generating level-specific descriptors that distinguish between competent and underperforming behaviors.

For ***communication and interpersonal skills***, excellent students communicated clearly and effectively, tailored explanations to patients’ understanding, used varied cueing strategies, provided comprehensive education that motivated engagement, built strong rapport, and contributed positively to the team. Good students communicated effectively enough to facilitate assessment and treatment, generally established rapport, and adapted communication in most situations but needed to simplify language and expand patient education. Adequate students gave simple, generally clear instructions and formed basic rapport, but frequently required clearer, more concise directions, more varied cueing, and better use of evidence when responding to professional questions. Not Adequate students showed unclear instructions, overuse of jargon, limited confidence or fluency, and difficulty adjusting language, pace, and volume to patients’ needs. A similar pattern was evident in documentation and presentation skills, with students rated as Excellent producing timely, detailed, accurate notes and confident, well-structured presentations and those at lower levels showing increasing problems with omissions, errors, poor organization, limited preparation and weak clinical understanding.

For ***clinical reasoning and decision making***, excellent students conducted logical, organized assessments, used appropriate outcome measures, integrated findings cohesively, and formulated realistic, patient-centered goals justified with hypotheses and evidence-based arguments. Good students generally used systematic frameworks (e.g. ICF), identified key problems, and set plausible goals, but needed refinement in prioritization and goal specificity and to avoid premature conclusions. Adequate students managed basic assessments and discharge plans, and recognized key problems but struggled to filter relevant information and link findings without guidance. Not Adequate students showed limited assessment structure and reasoning, weak knowledge of standard measures, and heavy reliance on educator prompts. The same pattern emerged in treatment planning and intervention: students rated as Excellent selected and progressed evidence-based interventions, monitored responses closely, and modified treatments creatively, whereas lower-rated students showed greater dependence on supervision, inconsistent outcome monitoring, limited and sometimes inappropriate exercise selection, and, at the lowest level, safety and organizational concerns.

For ***practical knowledge and technical competence***, Excellent students demonstrated precise, organized execution of manual techniques, familiarity with standardized tests and modalities, and safe, efficient handling, including complex transfers. Good students generally showed appropriate handling and competent techniques but required greater measurement accuracy and more practice with specific skills such as blood pressure measurement or transfers in frail older adults. Adequate students showed basic ability to perform systematic assessments and interviews, although with inaccuracies in measurement, occasionally rough or inefficient handling, and a need for closer supervision in choosing and progressing techniques. Not Adequate students frequently showed inaccurate range and strength assessments, poor palpation skills, unfamiliarity with key tests and equipment, and an inability to carry out many manual or modality-based interventions effectively without intensive guidance.

For ***learning attitude and reflective practice***, excellent students were consistently described as enthusiastic, proactive, and humble learners who actively sought feedback, asked questions, engaged with evidence, and used reflection to adjust their practice. Good students showed a positive attitude, participated actively in discussions and teaching activities, and generally implemented feedback, though were sometimes advised to be more consistently proactive and reflective. Adequate students were often eager and willing to accept feedback but showed variable initiative, weaker self-evaluation skills and a tendency for enthusiasm not always to translate into effective change. Not Adequate students demonstrated passivity, overconfidence, poor preparation, repeatedly forgot previously taught skills, and limited initiative to seek help or integrate feedback into clinical performance.

For ***professional behaviors and work ethics***, excellent students demonstrated strong reliability, consistent punctuality and attendance, safe and ethical practice, efficient time management across multiple cases, and mature, respectful interactions with patients and staff. Good students generally maintained high standards of professionalism, ethical behavior, and collaboration, but occasionally needed to improve punctuality, responsiveness to feedback, assertiveness, or time control. Adequate students showed caring and responsible behavior and overall satisfactory professional conduct, yet were sometimes undermined by distraction, low confidence, poor prioritization, or isolated unprofessional behaviors (for example, napping or inappropriate reactions). Not Adequate students exhibited repeated lateness and inadequate attendance, a weak sense of responsibility, difficulties maintaining professional boundaries and emotional regulation, and limited capacity to act independently and proactively in the clinical environment.

Finally, ***safety, risk management and patient-centered care*** showed a clear behavioral continuum. Excellent students displayed strong safety awareness, appropriate emergency responses, careful monitoring, good body mechanics, and highly patient-centered care characterized by sensitivity to pain, preferences, values and family context, with very positive patient feedback. Good students also practiced safely with minimal supervision and worked to keep patients comfortable and engaged but occasionally overlooked specific safety steps or their own ergonomics and were encouraged to adopt a more explicitly client-centered approach. Adequate students demonstrated basic safety awareness with observable improvement and caring, empathetic interactions, but had persistent gaps in recognizing red flags, avoiding unsafe acts, listening carefully to patient concerns, and maintaining appropriate physical distance. Not Adequate students showed recurrent safety lapses (for example, unsafe transfers, failure to monitor vital signs or heed contraindications, near misses with equipment), poor clinical judgment, and inefficient, uncomfortable assessment processes, requiring intensive supervision to prevent potential harm.

To illustrate how these attributes manifest across performance groups, Table 3 presents sample quotations for each GRS level mapped to the predefined attribute categories.

**Table 3.**
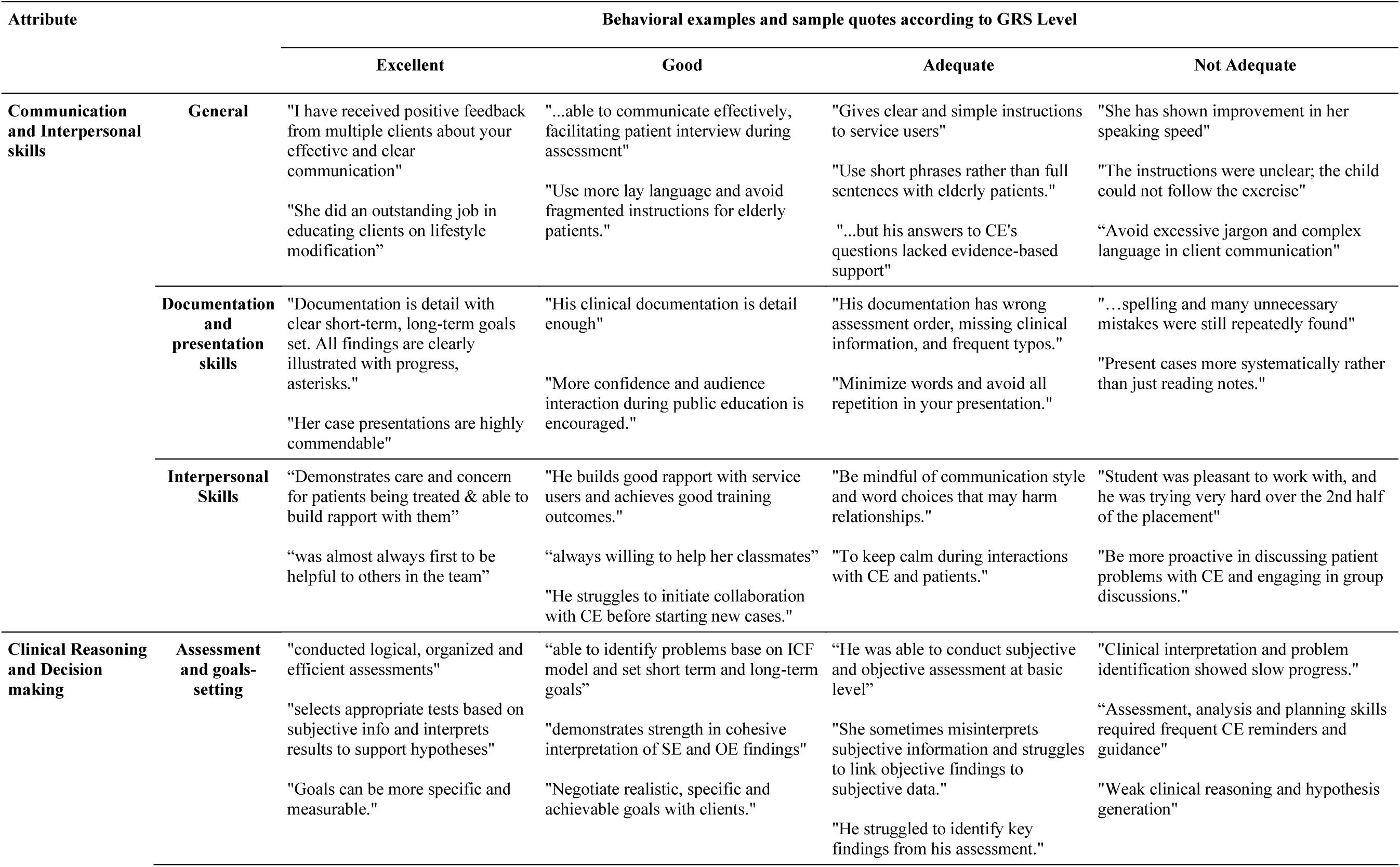
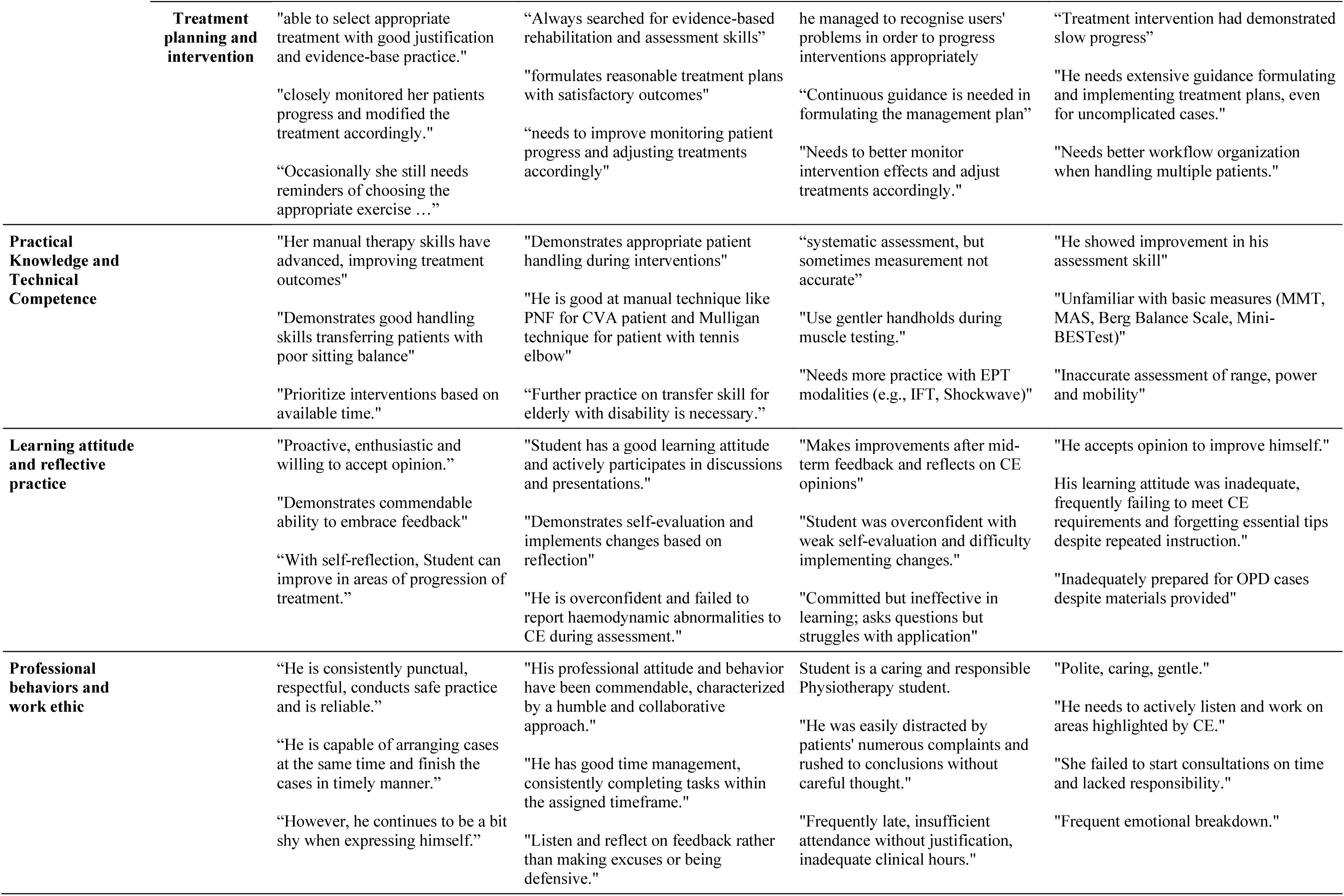
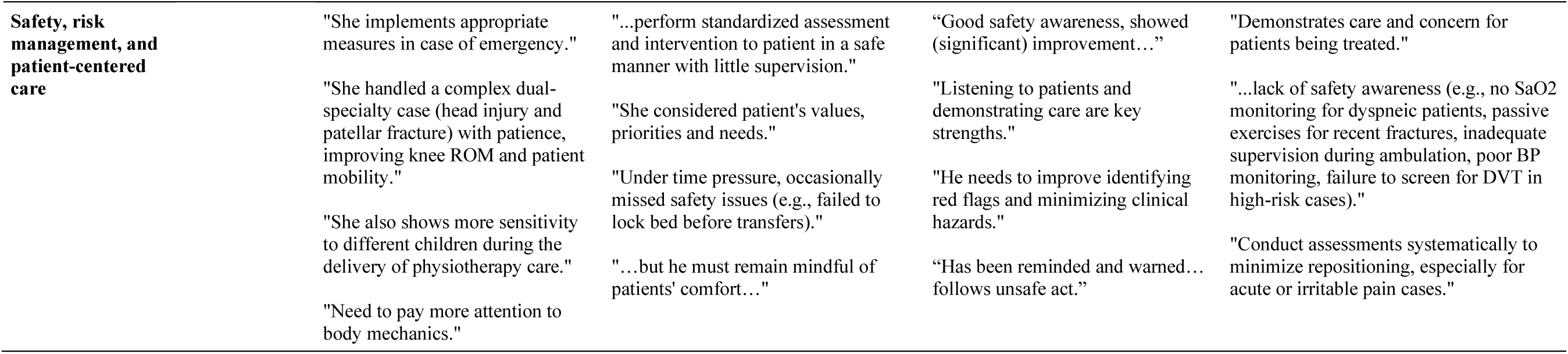
Sample quotes of student attributes across GRS performance levels.

## Discussion

This study is the first to explore the key attributes prioritized by HKCEs during physiotherapy placements and to describe the behavioral examples that distinguish performance levels on the Assessment of Physiotherapy Practice Global Rating Scale. From 456 qualitative comments by 45 CEs, six core student attributes were identified. While interprofessional competencies are common across disciplines, their interpretations vary, highlighting the need for precise behavioral descriptors (Brewer and Jones, 2013; John, Tong, Li, and Wilbur, 2020). These examples can help clarify performance expectations across competence levels and offer evidence-based guidance for students, educators, and curriculum developers. A key contribution of this work is addressing a persistent challenge in health profession education: assessment priorities are often implicit, and academic and clinical assessors may emphasize different aspects of performance than those stressed in classroom teaching, leaving students unsure how to reconcile theory with practice (Finstad, Knutstad, Havnes, and Sagbakken, 2022; Newton, Billett, Jolly, and Ockerby, 2009; O’Connor, McCurtin, Cantillon, and McGarr, 2018). While educators often share broad assessment criteria, they tend to prioritize and interpret these criteria differently when evaluating student achievement (Lipnevich, Mattern, and Feddock, 2025). By making explicit the standards and performance expectations that CEs use when assessing students, this study translates abstract competencies into practical, context-specific indicators that help bridge the theory–practice gap.

The Assessment of Physiotherapy Practice has demonstrated robust psychometric properties across diverse contexts and populations, supporting its utility as a reliable and valid measure of entry-level physiotherapy practice (Dalton, Davidson, and Keating, 2011, 2012). By analyzing the qualitative feedback from APP assessments in Hong Kong, this study identifies the behavioral indicators and performance standards that shape APP scores and shows how this established assessment framework is applied and understood within a specific cultural and educational context.

The six identified attributes align with existing competency frameworks for health professionals, but this study reveals how these competencies are prioritized in Hong Kong physiotherapy clinical practice (Canadian Interprofessional Health Collaborative, 2010; Frank, Snell, and Sherbino, 2015). Communication and interpersonal skills, clinical reasoning and decision-making, learning attitudes and reflective practice, and professionalism and work ethic emerged as the most consistently emphasized qualities, showing that CEs value not only what students know and can do, but also how they approach learning and professional growth. In the UK, the Chartered Society of Physiotherapy highlights communication, clinical reasoning, and professionalism among key graduate attributes, although lifelong learning is implied rather than explicit (Chartered Society of Physiotherapy, 2025). The emphasis on learning attitudes in the Hong Kong context very much aligns with the World Physiotherapy Education Framework, which identifies reflective practice and lifelong learning among eight core competency domains (World Physiotherapy, 2021). Furthermore, in this work, the strong emphasis on practical competence, followed by safety and risk management and patient-centered care, reflects expectations that physiotherapy education must address technical, cognitive, and affective dimensions to support safe, patient-centered practice (Berg-Carramusa, Mucha, Somers, and Piemonte, 2023).

### Learning attitudes and reflective practice: a particularly valued attribute in Hong Kong clinical education

The considerable specificity and frequency with which HKCEs documented learning attitudes and reflective behaviors in their feedback, including in both high and low GRS ratings, indicate that this competency is particularly valued in the Hong Kong context (Chong, Chau, Capio, and Jones, 2024). Students rated as excellent were described as enthusiastic, proactive, and humble learners who actively sought feedback, engaged critically with evidence, and used reflection to adjust their practice. Those rated as not adequate tended to be passive, overconfident, poorly prepared, and reluctant to seek help or integrate feedback. Health professions education literature increasingly recognizes reflective capacity and lifelong learning orientation as foundational to professional competence and clinical expertise, yet their consistent operationalization and assessment remain variable across contexts (Babenko et al, 2017; Fragkos, 2016; Or and Golba, 2024; Verkooijen et al, 2024). Reflective practice has been associated with critical thinking, self-awareness, clinical insight, quality of care, and ethical decision-making in complex cases, while engagement in learning is linked to professional identity, stress-related growth, and improved clinical competency (Khoshgoftar and Barkhordari-Sharifabad, 2023; Mitchell et al, 2023; Peng et al, 2025).

Comparative evidence suggests that HKCEs emphasize learning attitudes more strongly than in many other health professions education settings, where assessment often centers on knowledge, technical skills, and observable performance, with a less consistent focus on affective attributes such as learning attitudes and professionalism (Cotton, 2016; Landy et al, 2016; Ní Mhurchú and Cantillon, 2024). Reflexive practice is essential for shaping clinical judgment and improving outcomes, yet studies vary widely in how it is taught and assessed across disciplines (Landy et al, 2016). In Hong Kong, learning attitudes and reflective practice are consistently prioritized as core assessment domains rather than being treated as implicit or secondary expectations (Chong, Chau, Capio, and Jones, 2024). This strong focus in Hong Kong is closely tied to traditional Confucian values that shape ideas of good professional behavior. Medical professionalism in Hong Kong has been shown to draw on both Western ethics and Chinese thought, with ideals such as the “chun-tzu” (morally exemplary person) stressing virtue, humility, and constant self-improvement (Leung, Hsu, and Hui, 2012). Further, this may reflect cultural priorities, where values like continuous improvement, humility, and openness to feedback influence educator expectations and assessment (Leung, Hsu, and Hui, 2012). In East Asian contexts, these qualities are culturally significant and shape both educator approaches and student experiences (Chen, 2023; Reichenbach and Kwak, 2020). As such, this work’s documentation of learning attitudes as a core competency in the Hong Kong setting offers important insights for health profession education globally, particularly as educators increasingly recognize the need to align assessment with cultural and professional values (Walker et al, 2023).

### Methodological contributions

This study integrated inductive AI-assisted thematic analysis with deductive validation procedures and human interpretation, following new practices in qualitative health profession research methodology. AI tools facilitated the systematic processing of 456 participant comments, while researchers maintained oversight of all analytical stages, validating AI-generated codes, identifying contextually important variations requiring clinical judgment, and situating findings within physiotherapy education and practice (Cevik and Abu-Zidan, 2025; Hitch, 2024). This collaborative approach between human researchers and artificial intelligence takes advantage of the computational capacity of AI systems to identify patterns and thematic relationships while preserving the essential and irreplaceable contributions of human expertise in ensuring methodological rigor and research integrity (Cevik and Abu-Zidan, 2025; Hitch, 2024; Sakaguchi, Sakama, and Watari, 2025).

### Implications for Practice and Education

The identified attributes and behavioral examples provide curriculum developers with guidance for designing pre-clinical learning experiences that align with CEs’ expectations, provide CEs with structured frameworks for observing performance and providing consistent, justifiable Global Rating Scale scores, and offer students concrete benchmarks for self-assessment and targeted improvement during clinical placements. This aligns with contemporary frameworks emphasizing feedback literacy, a learner’s ability to recognize, comprehend, and act on feedback to aid in the development of clinical competency (Tripodi, Feehan, Wospil, and Vaughan, 2021). By providing concrete behavioral examples characterizing each Global Rating Scale level, this study supports the development of a shared understanding of performance standards among stakeholders, which is essential for effective feedback, valid assessment, and meaningful learning (Pangaro and Ten Cate, 2013; Williams et al, 2024).

This study’s systematic analysis of CE feedback aligns with contemporary competency-based education frameworks that emphasize observable behaviors and clear competency descriptions (Al Jabri, Kvist, Azimirad, and Turunen, 2021). By grounding these attributes in the actual language and behaviors of experienced CEs rather than relying solely on theory or expert consensus, this study enhances the authenticity and contextual relevance of its competency descriptors. This evidence-based, locally contextualized approach bridges abstract competency frameworks with the real-world assessment practices of clinicians.

### Study Limitations

The analysis was limited to written qualitative comments in the final feedback section of the APP forms, so the identified attributes reflect only those aspects of student performance that the CEs considered sufficiently important to document in writing. A second consideration relates to the program-specific context: the study involved one Hong Kong physiotherapy program, so the findings may reflect local educational and clinical priorities and may differ from those in other geographical or institutional settings. However, the dataset comprising 456 feedback comments from 45 CEs across multiple specialties and practice settings provides substantive depth and diversity within the Hong Kong context. The six identified attributes align with systematic review findings on core competency frameworks, which highlight domains such as professionalism, ethical practice, research, personal development, teamwork, leadership, and patient-centered care (Musolino and Jensen, 2024). This alignment supports the broader applicability of the findings while recognizing HKCEs’ distinctive emphasis on learning attitudes and reflective practice.

### Conclusion

This study identified the key attributes prioritized by HKCEs and characterized the behavioral examples that distinguish performance levels in the Assessment of Physiotherapy Practice. By systematically analyzing CE feedback, this study reveals the implicit competency standards and performance expectations embedded in clinical assessment data. A distinctive finding is the prominence HKCEs assign to learning attitudes and reflective practice as core competencies, rooted in traditional Confucian values of continuous self-improvement, humility, and professional duty. The identified attributes and behavioral descriptors provide practical resources for improving alignment between academics and CEs, clarifying expectations for students, and supporting more consistent and equitable assessment practices in physiotherapy clinical education. Future studies should explore how using these findings in curriculum planning, clinical educator training, and pre-clinical preparation affects feedback quality, assessment decisions, and students’ readiness for independent practice.

## Data Availability

All data produced in the present study are available upon reasonable request to the authors.

## Acknowledgments

This work was supported by the [Teaching and Learning Research Fund of Hong Kong Metropolitan University] under grant [Reference number TLRF-2024/05].

Declaration of interest

The authors declare no conflicts of interest.

## Ethics Approval

Ethical approval was obtained from the Hong Kong Metropolitan University Research Ethics Committee (REC) (Reference number: HE-T&L2024/05, Date of approval: 23 December 2024).

## Informed consent and patient details statement

No separate Personal Information Collection Statement or informed consent was required for this secondary analysis, as the course scores, practicum scores, and admission information were originally collected as part of routine academic administration and program management. All the data used in this research were de-identified prior to analysis, and no individual student could be identified from the results presented.

